# Feasibility assessment of LI-RADS based ultrasonic screening and CT diagnosis for hepatocellular carcinoma in low middle income setting: observation from a small study

**DOI:** 10.1101/2023.11.08.23297434

**Authors:** Ephranzia Chao Malindi, Timothy Musila Mutala, Stephen Onyango Ayo, Roseline Kerubo Ogaro

## Abstract

In our cross-sectional study, we assessed the feasibility of using the ACR LI-RADS screening and diagnostic pathway for Hepatocellular Carcinoma (HCC) in a low-middle-income setting. HCC develops in a well-defined high-risk population, but many countries in this income category lack HCC screening and surveillance guidelines. The ACR’s practical approach through LI-RADS provides a valuable solution. Our study showed that 5.66% of patients had positive ultrasound findings, and significant lesions were detected on multiphasic CT scans. Out of these, 1.89% were diagnosed with definite HCC (LR-5 lesions). We conclude that HCC screening and diagnosis using ACR LI-RADS algorithms are feasible in such settings. However, success hinges on knowledge, skills, and technical capacity building, and it is vital to consider the downstream effects of the screening and surveillance program during the design process.

**Summary:** HCC screening and diagnosis using the ACR LI-RADS algorithms are feasible in low-middle-income settings, making it applicable and recommendable for even lower-income countries. However, successful implementation requires a focus on knowledge, skills, and technical capacity building.

**Key points:**

- Eligible patients for hepatocellular carcinoma screening and surveillance programs are easily identifiable in low middle income countries.
- American College of Radiology’s Liver Imaging Reporting and Data Systems algorithm provides a practical approach to hepatocellular carcinoma screening and surveillance.
- With proper knowledge, skills, and technical capacity, this algorithm is feasible in a low middle income setting, factoring tiered levels of care and referral systems.

## Introduction

Globally, liver cancer is the sixth in terms of incidence and the third in mortality causation of all cancer sites as per GLOBOCAN 2020 estimates, translating to approximately 906, 000 and 830,000 new cases and deaths respectively (1). Our country is a low middle income country within Eastern Africa in which the age-standardized incidence of liver cancer is 6.2 per 100,000 for males and 4.0 per 100,000 for females as per the same report. The specifics for the country indicate that there are 924 new cases and 856 deaths annually from the disease in the detailed GLOBOCAN 2020 report (2). Hepatocellular carcinoma (HCC) develops in a well described high risk population that is defined by viral hepatitis, alcoholic cirrhosis, non-alcoholic liver cirrhosis from obesity and diabetes and aflatoxin as major causes (3). Our tertiary hospital, the largest within the Eastern and Central Africa region runs a dedicated liver clinic where patients with these well identified risk factors are followed up. We noted with concern that the latest cancer screening guidelines in our country developed in 2019 did not include HCC (4). We infer that HCC screening satisfies the Wilson and Jungner’s standards and principles of screening (5) in our local situation, given the above epidemiological briefs. In addition, there has been a capacity building development on possibility of early treatment of HCC within our tertiary hospital. A team of surgeons have been trained in hepatobiliary surgical subspecialty and from our experience they perform about 20 liver resections in a year. We also have a team of qualified interventional radiologists that have been administering local treatment for HCC as per indications, and we have fellows undergoing training which further builds the capacity. An icing on the cake is that our tertiary hospital has a functional hepatobiliary multidisciplinary team (MDT) meeting every Tuesday that deliberates on treatment options for patients.

Multiple guidelines have recommended imaging and/or alpha-fetoprotein as a screening method for HCC in high risk groups of patients. These include American Association for the Study of Liver Diseases (AASLD), National Comprehensive Cancer Network (NCCN), European Association for the Study of Liver (EASL) and Japanese Society of Hepatology (JSH) among others. The American College of Radiology (ACR) has provided a practical approach to the imaging in HCC screening through the Liver Imaging and Reporting Data Systems (LI-RADS). Recognizing that ultrasound (US) is the mainstay imaging modality for the task, US LI-RADS was developed with components of visualization and detection scores (6). The detection scores are divided into three categories: US-1(negative), US-2 (subthreshold) and US-3 (positive). Positivity under the US LI-RADS algorithm is favorably comparable with other previously established guidelines like AASLD and EASL in that a suspicious nodular lesion is one of longest diameter >1 cm (7,8). The US-3 (positive) lesion is subjected to multiphasic imaging, mainly computed tomography (CT) or magnetic resonance imaging (MRI) whereby diagnostic CT/MRI LI-RADS characterization is performed (9). Contrast enhanced ultrasound (CEUS) has also been incorporated as an emerging tool in the diagnostic LI-RADS category assignment. In our setting US for the initial screening process is ubiquitously available and relatively cheap. Access to multidetector CT (MDCT) scans that meet the technical requirements for LI-RADS diagnostic workup is well assured in most of the referral hospitals within the country. MRI machines of 1.5T and 3.0T magnetic strength are far much fewer and mainly concentrated within the capital city. CEUS technology has not been established in our country.

It is against the above-described background that we set out to assess the feasibility of using the ACR LI-RADS screening and diagnostic pathway with defined endpoint outcomes as the number of positive US nodules, CT LI-RADS categories from the positive sonographic lesions and the proportion of definite HCC (LR-5) lesions.

## Material and Methods

This was a cross-sectional study designed to pass patients presenting to the liver clinic through a LI-RADS based US screening and CT diagnosis whenever indicated between July 2020 and January 2021. Following our local (KNH-UON Ethics & Research Committee) institutional ethical review committee approval (P1029/12/2019) and a small local hospital grant, 150 patients were consecutively sampled for the study. This was way above the desirable sample size of 102 based on Fischer’s formula for participants with the condition of interest and finite population definition. Written formal consent was obtained from the participants. The inclusion criteria were cirrhosis from any cause and patients on follow up for hepatitis B and C infection. Excluded were patients with previously diagnosed HCC or other primary malignancy site, participants who did not provide informed consent, those unable to undergo the entire process of the evaluation and where adequate relevant data was unavailable.

The included participants underwent liver ultrasound protocol as published by experts panel for US LI-RADS (6), covering the technical and interpretation aspects. Sonographers allocated to the study were sensitized to the required protocol for the study and the two radiologists who were investigators in the study also performed some of the sonograms. Each sonographic examination outcome was allocated a US LI-RADS visualization and detection score through consensus and review by the researchers. Positive (US 3) lesions were subjected to multiphasic 128 slice multi-detector CT scanner at 120 Kv, collimation of 1.5mm and a pitch of 1.88. The applied phases were pre contrast, arterial phase (35s), porto-venous phase (75s) and a delayed phase (120s), all done at inspiratory breath-hold. The findings from the CT multiphasic imaging findings led to assignment of a LI-RADS diagnostic category to each of the US-3 lesions. The number of lesions categorized in each stage of the pathway was determined and tabulated with proportions in relation to the sample size. Negative (US-1) and subthreshold (US-2) outcomes were referred to routine 6 monthly surveillance and 3 monthly follow up, respectively. All the positive cases were referred for MDT meeting.

## Results

Out of 150 identified eligible patients, 106 met the inclusion criteria which became our sample size. Simple demographic descriptors showed 65 (61.3%) male and 46 (38.7%) female patients of age range 20-82 years (median 37.5).

All the US examinations were of minimal limitation visualization score (A). out of the 106, six (5.66%) were US-3, all of which underwent multiphasic CT examination.

The CT LI-RADS categories for the lesions were as follows: LR-3 one, LR-4 one, LR-5 two, LR-M, one and LR-TIV one (table 1).

**Table 1:** Number of lesions in each CT LI-RADS defined categories.

| **LI-RADS Category** | **Number** |
| --- | --- |
| LR-NC | 0 |
| LR-1 | 0 |
| LR-2 | 0 |
| LR-3 | 1 |
| LR-4 | 1 |
| LR-5 | 2 |
| LR-M | 1 |
| LR-TIV | 1 |
| **Total** | **6** |

The participants’ flowchart is presented in figure 1.

**Figure 1:**
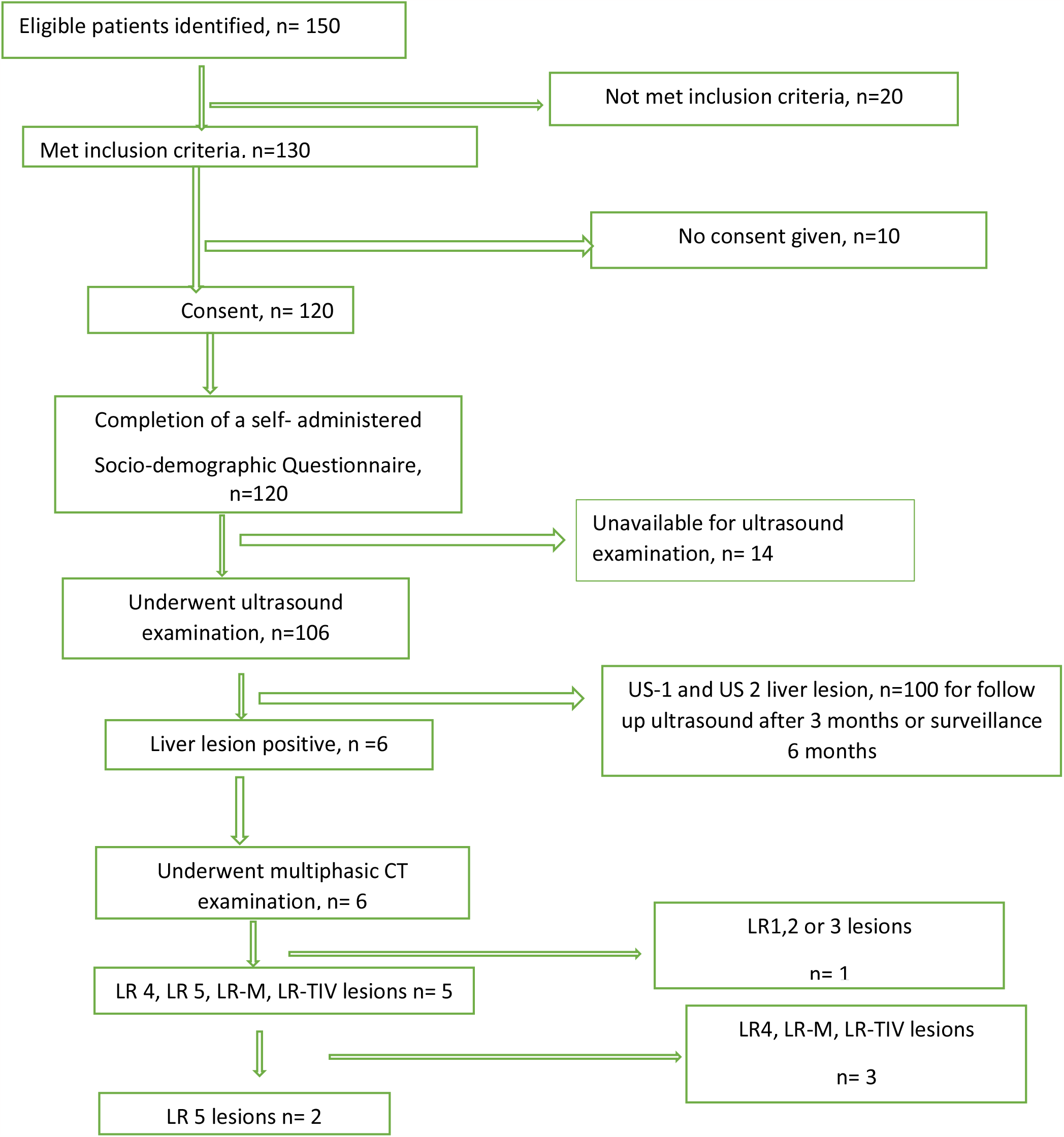
Participants’ flowchart.

Overall, the percentage of the LR-5 lesions that represents definite HCC was 1.89% in this single point full run of the combined US and CT LI-RADS assessment.

Figures 2,3 and 4 demonstrate representative US and CT images from our study.

**Figure 2:**
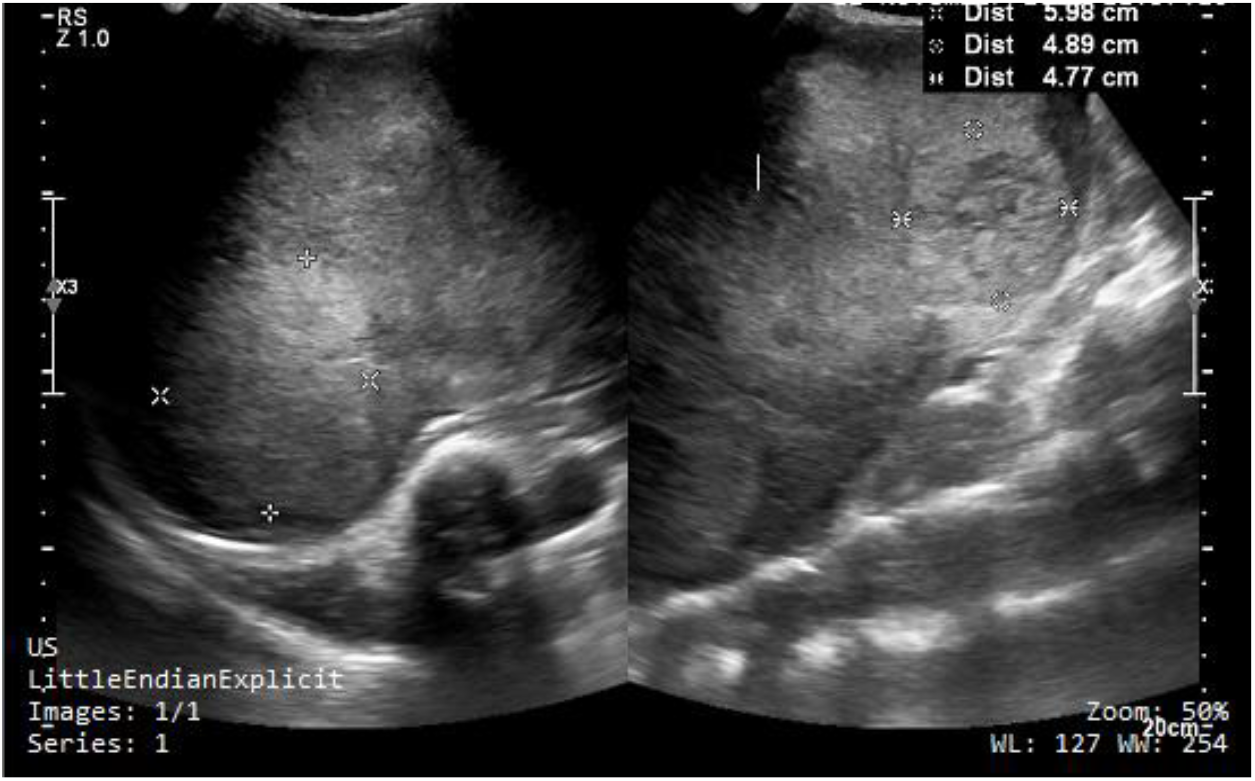
Gray scale image of the liver demonstrating multiple lobulated hyper echoic lesion.

**Figure 3:**
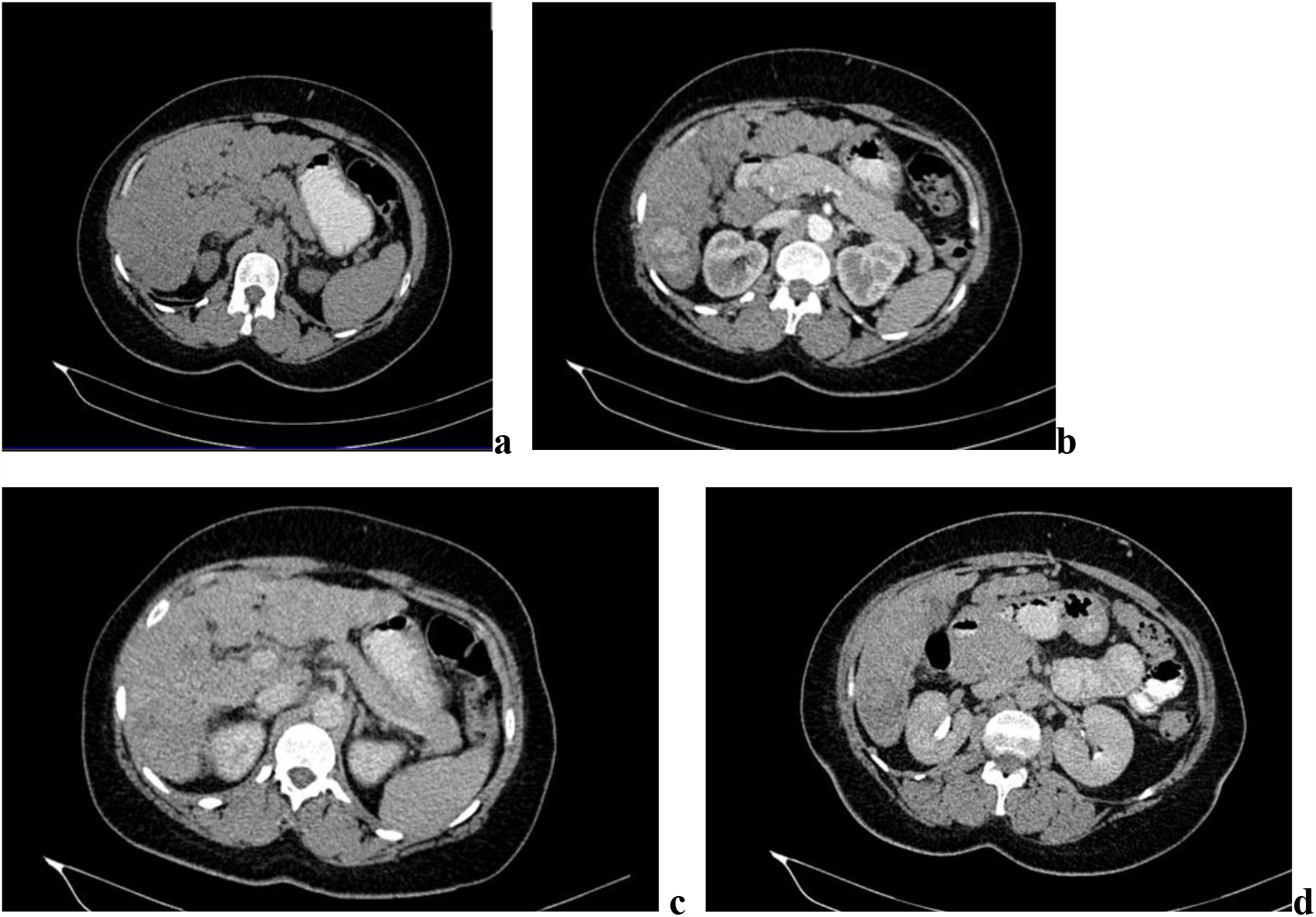
**A 3.96 cm lesion within segment six which is isodense to the liver precontrast (a), non-peripheral arterial hyperenhancement (b), non-peripheral porto-venous washout (c) and hypo attenuating in delayed phase (d). all features of an LR 5 lesion**.

**Figure 4:**
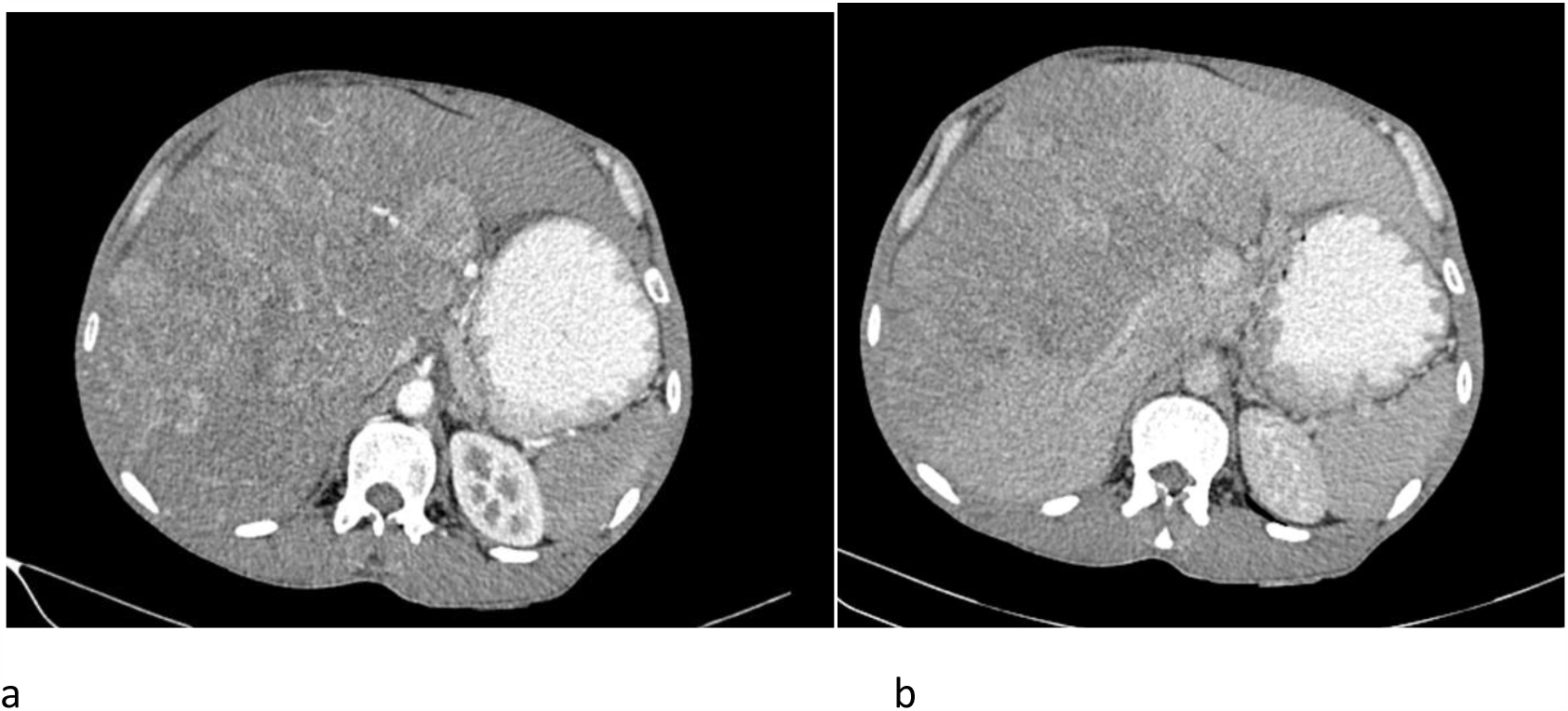
**An LR- TIV Multi-nodular lesion involving both hepatic lobes with arterial hyperenhancement (a) and porto-venous washout (b). There was hepatic artery involvement and other cuts (not shown) demonstrated tumor-in-vein**

## Discussion

After establishing that HCC was of public health concern in our community and that our national cancer screening guidelines omitted it and knowing that there is a radiological approach to effecting it, we set out to provide leadership in that aspect. We successfully ran our sample through the entire process of screening and diagnosis applying the ACR-LI-RADS criteria. It is not only our country within Sub-Saharan Africa that lacks national policies on HCC screening and surveillance but the conversation on this matter is catching up (10,11). For national policy development on matters that touch on screening, the capacity must be addressed. We found the ACR-LI-RADS algorithm suitable for testing the capacity and performance of HCC screening. We can propose it as a standard for other low and low-middle income countries, especially within Sub-Saharan Africa. The equipment and performance technical specifications are well outlined in the ACR website (12). Radiology must play a central and leading role in initiating the screening and surveillance of at risk patients for this type of cancer. Evidence continues to build up that screening and surveillance improves outcomes in HCC (14). At the same time, there must exist an MDT approach for such a program to ensure that outcomes are optimized for the most appropriate patient care. The levels of care may be structured differently across countries but in our country, it is easy to allocate roles and responsibilities based on the already mapped out resources at each level of care.

Though our feasibility study was in a tertiary care setting (designated as a level 6 facility in our local grading), we can figure out that at least all level 4 county referral hospitals can carry out the US screening and surveillance tasks. The diagnostic arm can be taken up by level 5 hospitals since they are equipped with MDCT scanners. Any private facilities with assessed capability to perform both tasks can also be brought on board as well. Of course, all this will depend on training that is standardized with the ACR LI-RADS.

Our small study of single run screening without surveillance returned a yield of 5.66% pickup rate of suspicious nodules on ultrasound, which also showed significant findings on MDCT. Another pilot study in Zambia, another Sub-Saharan Africa country of low-income status reported a 2% rate, though their focus was only on HBV-HIV co-infection and had a larger sample size of 279 patients (10). A larger surveillance study in USA found that 25% of patients in the program had suspicious sonographic lesions that warranted further workup (13). The downstream effects of HCC screen surveillance is still a matter of investigation.

Our small study is limited by the fact that we had only a single point screening for the patients therefore sweeping conclusions on the prevalence and incidence cannot be derived from the same setting as a continuous surveillance program. Also external validation within our peripheral health facilities was not carried out, a necessary step after training the key providers in its use.

## Conclusions

HCC screening and diagnosis is feasible using the ACR LI-RADS algorithms in a low middle income setting which we can recommend for even lower income countries. This must be matched with knowledge, skills, and technical capacity building. Establishing the downstream effects of the screening and surveillance program should be at the core of the design.

## Supporting information

ERC approval

## Data Availability

none

## Abbreviations

HCC: hepatocellular carcinoma
US: ultrasound
ACR: American College of Radiology
LI-RADS: Liver Imaging and Reporting Data Systems
CT: Computed Tomography
GLOBOCAN: Global Cancer Observatory
MDT: multi-disciplinary team
AASLD: American Association for the Study of Liver Diseases
NCCN: National Comprehensive Cancer Network
EASL: European Association for the Study of Liver
JSH: Japanese Society of Hepatology
CEUS: contrast enhanced ultrasound
MRI: magnetic resonance imaging
HBV: hepatitis B virus
HIV: human immunodeficiency virus
KNH: Kenyatta National Hospital
UON: University of Nairobi
ECM: Ephranzia Chao Malindi
TMM: Timothy Musila Mutala
SOA: Stephen Onyango Ayo
RKO: Roseline Kerubo Ogaro

## Acknowledgments

We sincerely thank Dr Gladys Mwango then Chairperson Department of Diagnostic Imaging and Radiation Medicine, University of Nairobi, and Dr Mamai Amo Head of Department Radiology, Kenyatta National Hospital (KNH) for their human resource and material support.

We also acknowledge Mr. Wycliffe Ayieko our statistician for helping in determining the sample size. We are grateful to KNH Research Program for the grant to perform this work. Lastly, we sincerely recognize the efforts of Miss Veronicah Wanjiku Kung’u in data management for our study.

## Author contributions

All authors contributed equally to the study conception. SOA contributed to identifying the relevant patients for the study in the liver clinic. ECM, RKO and TMM contributed to the performance of the radiological examinations and interpretation. All authors contributed equally to the manuscript preparation.

## Disclosures to conflicts of interest

None of the authors declares any conflict of interest

