## Supplementary material for "Feasibility assessment of LI-RADS based ultrasonic screening and CT diagnosis for hepatocellular carcinoma in low middle income setting: observation from a small study": ERC approval

UNIVERSITY OF NAIROBI
COLLEGE OF HEALTH SCIENCES
P O BOX 19676 Code 00202
Telegrams: varsity

KNH-UON ERC

Website: http://www.erc.uonbi.ac.ke
Facebook: https://www.facebook.com/uonknh.erc
Twitter: @UONKNH_ERC https://twitter.com/UONKNH_ERC

KENYATTA NATIONAL HOSPITAL
P O BOX 20723 Code 00202
Fax: 725272
Telegrams: MEDSUP, Nairobi

Ref: KNH-ERC/A/146

14th May 2020

Dr. Ephranzia Chao Malindi
Reg. No.H58/8190/2017
Dept. of Diagnostic Imaging and Rad. Medicine
School of Medicine
College of Health Sciences
University of Nairobi

Dear Dr. Malindi

**RESEARCH PROPOSAL – ULTRASOUND IMAGING DETECTION OF HEPATOCELLULAR SUSPICIOUS LESION AMONG HIGH RISK PATIENTS ATTENDING KENYATTA NATIONAL REFERRAL HOSPITAL LIVER CLINIC (P1029/12/2019)**

This is to inform you that the KNH- UoN Ethics & Research Committee (KNH- UoN ERC) has reviewed and **approved** your above research proposal. The approval period is 14th May 2020 – 13th May 2021.

This approval is subject to compliance with the following requirements:

a. Only approved documents (informed consents, study instruments, advertising materials etc) will be used.
b. All changes (amendments, deviations, violations etc.) are submitted for review and approval by KNH-UoN ERC before implementation.
c. Death and life threatening problems and serious adverse events (SAEs) or unexpected adverse events whether related or unrelated to the study must be reported to the KNH-UoN ERC within 72 hours of notification.
d. Any changes, anticipated or otherwise that may increase the risks or affect safety or welfare of study participants and others or affect the integrity of the research must be reported to KNH- UoN ERC within 72 hours.
e. Clearance for export of biological specimens must be obtained from KNH- UoN ERC for each batch of shipment.
f. Submission of a request for renewal of approval at least 60 days prior to expiry of the approval period. (*Attach a comprehensive progress report to support the renewal*).
g. Submission of an *executive summary* report within 90 days upon completion of the study. This information will form part of the data base that will be consulted in future when processing related research studies so as to minimize chances of study duplication and/ or plagiarism.

For more details consult the KNH- UoN ERC website**http://www.erc.uonbi.ac.ke**

Yours sincerely,

**PROF. M. L. CHINDIA**
**SECRETARY, KNH-UoN ERC**

c.c. The Principal, College of Health Sciences, UoN
The Director, CS, KNH
The Chairperson, KNH- UoN ERC
The Assistant Director, Health Information, KNH
The Dean, School of Medicine, UoN
The Chair, Dept. of Diagnostic Imaging and Radiation Medicine, UON
Supervisors: Dr. Mutala MusilaTimothy, Dept.of Diagnostic Imaging and Radiation Medicine, UoN
Dr. Onyango Stephen Ayo, Dept.of Internal Medicine, KNH
